# Epidemiology and clinical profile of sports injuries treated in the Douala General Hospital and the Douala Laquintinie Hospital

**DOI:** 10.1101/2023.09.15.23295614

**Authors:** Tankeng Leonard Tanko, Ngatchou Djomo William, Theophile Chunteng Nana, Mbianyor Bill-Erich, Akwa Gilbert, Lifafa Kinge Kange, Aimé Bonny

**Author notes:** Corresponding author (TLT). These authors contributed equally to this work. Current Address: Emergency Department, Hopital Saint Jean de Malte, Njombe, Littoral, Cameroon (Tankeng Leonard Tanko). Current Address: Department of Surgery, Douala General Hospital, Littoral, Cameroon (Ngatchou Djomo William).

## Abstract

**Background:** A sport injury is any body tissue damage that results from the practice of sports or exercise. Despite the number of cases treated at hospitals, very little is reported. Regarding the rising participation in sports and as a consequence sports injuries, this study aimed to present the epidemiology and clinical profile of sport injuries treated at two tertiary hospitals in Douala Cameroon.

**Methods:** This was a hospital based cross sectional descriptive study during nine months, carried out in the Douala General hospital and the Douala Laquintinie Hospital. Medical records of patients who were treated for sports injuries from January 2012 to April 2022 were included in the study. Severe injury was considered to be an injury score of greater than or equal to three according to the abbreviated injury scale of the injured body region.

**Results:** Among 72 consecutive eligible records, there was a male predominance (86.1%) with a 6.2 M:F sex ratio. The mean age 24.97±13.8 years and the modal age range was 11-21 years. The prevalence of sports injuries was 1.12% amongst all trauma cases. 48.6% of injured persons were students, 36.1 % employed, 8.3% athletes and 6.9% unemployed. 59.7% of injuries occurred during recreational sports while 22.2% occurred during physical education, 11.1% during trainings and 6.9% during competitions. Football accounted for 66.7% injuries, running 13.9% and 12.5% in jumping. 59.7% of injuries were fractures, 6.9% dislocations, 27.7% soft tissue injuries and 5.6% head injuries. 47.2% of injuries occurred on the lower extremities, 29.2% on the upper extremities, 6.9% on the face, 5.6% abdominopelvic, 4.2% on the thorax, 5.6% head injuries and 1.4% on the neck. Overall 73.7% of sports injuries were severe injuries.

**Conclusion:** Sports injuries treated in hospitals are severe injuries. The majority of sports injuries treated in hospitals are fractures. Most sports injuries occur in the lower extremity. Football records the highest number of injuries.

## Introduction

Sport refers to all types of physical activity which, through random or organized participation, are directed towards the improvement or expression of physical and mental well-being, achievement of results in competitions of all levels or for the formation of social relations [1]. The practice of sports develops physiological and psychosocial benefits [2]. However, the practice of sports is inevitably linked with the occurrence of injuries [3]. A sports injury is defined as body tissue damage that occurs as a result of sport or exercise. The European Sport Charter (ESC) considers any body tissue damage during physical activity as a sport injury. Physical activity includes numerous forms of activity such as working, fitness exercise, outdoor activity, playing, training, getting in shape, working out, and physical education. Sport injuries can be divided into acute injuries and overuse injuries [4].

According to a study carried out during the PyeongChang 2018 Olympic Winter Games Overall, 12% of the athletes incurred at least one injury during the Games [5]. In the United States, it was reported that 3.5 million youth under the age of 15 years received medical care each year for injuries that occurred during sports practice with two-thirds of those injuries requiring care in emergency units [6]. In Liverpool, amongst 2432 new patients seen in an accident and emergency department 7.1% were sports related [7].

Akkaya et al, in an analysis of sport related injuries at an emergency department in Turkey showed that 80.5% of cases were men, and the mean age was 25.7±5.1 years. Football injuries were a majority with 73.1% cases, then basketball injuries in 14.0% cases, and running or walking injuries in 10.2% cases. Football was the most common cause of injury identified in all age groups, basketball was the second cause of injury for age under 35 years. Running and walking was the second cause of injury in those over 35 years. The most commonly injured body region was the lower extremities 62.7%, followed by the upper extremities 23.3% and the head and neck 6.9% respectively. While strains and sprains were more frequent in basketball, fractures-dislocations and other superficial injuries were more common in artificial surface football games [8].

In Cameroon, a community based study done in Yaoundé amongst athletes in six sports (karate, judo, basketball, handball, football and wrestling) reported a prevalence of 47.9% and 37.1% for bodily and dentofacial injuries respectively. The distribution of the bodily injuries were limbs 60.0%, chest 23.5%, abdomen 11.3% and neck 5.2% [9]. The increasing rate of population participation in physical activity, amateur and professional sports is linked with the occurrence of injuries. Sport injuries treated in hospitals are often severe with long lasting musculoskeletal and psychosocial consequences to the injured individuals or athletes. This has motivated us to study the epidemiological aspects and the clinical aspects of sports injuries treated in hospitals. This study will provide us with the characteristics of sports injuries treated at the hospital, the different anatomical regions concerned, the severity of these injuries and the types of sports involved in our context.

## Methods

### Study Characteristics

This was a hospital based, cross-sectional descriptive study, carried out from 1^st^ October 2021 to 30^th^ June 2022 a duration of nine months. We studied medical records of patients who were treated for sports injuries from January 2012 to April 2022 a period of nine years four months. This study was carried out at the Douala General Hospital (DGH) and the Douala Laquintinie Hospital (DLH). These two hospitals are reference hospitals in the city of Douala which receive professionals, amateurs and non-athletic regular persons who participate in recreational, national and international sports activities. Douala is the capital city of the Littoral Region of Cameroon, a country in sub-Saharan Africa. It is the largest city and the economic capital of the country, with a population of more than 3.7 million inhabitants [10].

### Study Population and Sampling

The study targeted file records of individuals presenting any injury which occurred during sports or exercise at the Emergency Department (ED) or Out Patient Department (OPD) of the DGH and DLH. Sampling was consecutive. All file records at the emergency and outpatient departments were screened for eligibility. Inclusion criteria were: All file records of patients treated for any body tissue damage during sports and exercise in the period of study at DGH and DLH. Exclusion criteria included all incomplete file records.

### Ethical Considerations

The study was conducted only after ethical and administrative clearance had been obtained from the University of Douala (No.2987 IEC-UD/04/2022/T), the Review Board of DGH (N°_AR/MINSANTE/HGD/DM/03/22) and the DLH (N° 01077/AR/MINSANTE/DHL). The confidentiality of patients was maintained by using serial numbers rather than names on data extraction form. Data was obtained following recommended guidelines and coded to ensure anonymity.

### Study Procedure

After obtaining ethical clearance and administrative approvals, patient files records that met the inclusion criteria were sorted out from the archives of the OPD and the ED at the DGH and DLH. File records of patients who were hospitalised and managed after consultation at the OPD or ED were further sorted out at the corresponding hospitalisation services. Clinical data extracted was coded and entered into a data extraction form then transferred into a data collection software.

### Statistical Analysis and Definitions

The data collected was coded and input into Open data kit collect (ODK Collect) version 2022.1.2 and exported to Microsoft Excel 2016 where data was managed, cleaned then analysed with SPSS version 25.0. Quantitative variables were presented as mean (and standard deviation), or as frequencies and percentages. Results were represented on tables and figures (pie charts, bar charts, histograms) to ease organization and comprehension. Collected data were sociodemographic characteristics (age, sex, occupation), Skill level (Amateur, Professional, Non athletic regular person), Reason of participation (Recreational, Training, Competition, Physical Education), Type of sport or exercise practiced (Football, Handball, Walking, Running, Volleyball, Basketball, Tennis, swimming, cycling, Aerobics, Weight training, Wrestling, Boxing, Judo, Jumping, Karate, Dance, Other) Injury mechanism: Direct contact (athlete, object, other), Indirect contact, no contact, unknown mechanism, Soft tissue injury; (Sprain, Contusion, Abrasion, Laceration, Strain, Hematoma, Other), Hard Tissue injury; Fracture, Dislocation, Other, Anatomical region concerned (Head, Neck, Face, Thorax, Abdomen, Pelvis), Anatomical region concerned: (Upper Extremities; Shoulder, Arm, Elbow, Forearm, Wrist, Hand) Anatomical region concerned: (Lower Extremities: Hip, Thigh, Knee Leg, Ankle, Foot), Injury Severity (AIS; 1 Minor 2 Moderate 3 Serious 4 Severe 5 Critical 6 Maximum). Severe sport injuries were defined as any sport injury with a score of greater than or equal to three (3) on the abbreviated injury scale of the concerned body segment. Indirect contact injuries were injuries that occurred in a body region different from the site of impact or collision. Non-athletic regular person was defined as any individual without physical disability who practises sport mainly for recreational and fitness purposes, or does not practise sport at all.

## Results

### Sociodemographic Characteristics

A total of 10,211 file records of patients who reported to the outpatient and emergency department for trauma were reviewed, 114 were treated for sports injuries a hospital prevalence of 1.12% amongst which 42 were excluded due to incomplete records. A total of 72 file records were retained in our study. There was a male predominance with 62 (86.1%) cases and 10 females (13.9%), giving a M:F sex ratio of 6.2. The modal age range was 11-21 years, the mean age was 24.97±13.8 years. The ages ranged from 3 to 69 years old. Students were found to have the highest sports injuries with 35 cases (48.6%), the employed with 26 cases (36.1%), 6.9% unemployed and athletes with 6 cases (8.3%). 88.9% of injured individuals were non-athletic regular persons, 9.7% were amateurs and 1.4% were professionals. Recreational sports or exercise accounted for 43 cases (59.7%) while 22.2 % of injuries occurred during physical education, 11.1% occurred training and 6.9% during competitions. The majority of injuries occurred during football 48 (66.7%).

**Table I:** Type of sport or exercise practiced by patients treated for sports injuries at the DGH and DLH during the period of study.

| Variable | Frequency (n=72) | Percentage (%) |
| --- | --- | --- |
| Football | 48 | 66.7 |
| Running | 10 | 13.9 |
| Jumping | 9 | 12.5 |
| Basketball | 2 | 2.8 |
| Dance | 1 | 1.4 |
| Karate | 1 | 1.4 |
| Tennis | 1 | 1.4 |
| Total | 72 | 100.0 |

### Clinical Characteristics

43 cases (59.7%) were direct contact injuries; whereby 25 cases was contact with athletes, 9 with objects (balls, goal, bats) and 9 with play surface. 21 cases (29.2%) were indirect contact injuries and 8 no contact injuries (11.1%). 66.6% of injuries were hard tissue injuries with fractures having 43 cases (59.7%) and dislocations 5 cases (6.9%). Soft tissue injuries represented 27.7% with sprains being the highest soft tissue injury. 5.6% were head injuries.

**Table II:** Type of sports injuries treated at the DGH and DLH during the period of study.

| Variable | Frequency (n=72) | Percentage (%) |
| --- | --- | --- |
| **Hard Tissue Injury** |  |  |
| Fractures | 43 | 59.7 |
| Dislocation | 5 | 6.9 |
| **Soft Tissue Injury** |  |  |
| Sprain | 6 | 8.3 |
| Contusion | 5 | 6.9 |
| Strain | 5 | 6.9 |
| Laceration | 4 | 5.6 |
| **Head Injury** | 4 | 5.6 |
| Total | 72 | 100.0 |

Most of injuries occurred on the lower extremity (47.2%) and 29.2% on the upper extremity. Among 34 injuries that occurred to the lower extremity; 11 cases occurred on the ankle, 10 cases on the knee, 2 cases on the hip, 2 cases on thigh, 9 cases on the leg. In the Upper extremity 21 injuries were recorded with 13 occurring on the forearm, 4 on the shoulder, 3 on the elbow and 1 on the wrist.

**Table III:** Anatomical distribution of sports injuries in patients treated for sport injuries at DGH and DLH during our period.

| Variable | Frequency (n=72) | Percentage (%) |
| --- | --- | --- |
| Lower Extremity | 34 | 47.2 |
| Upper Extremity | 21 | 29.2 |
| Face | 5 | 6.9 |
| Abdomen and pelvis | 4 | 5.6 |
| Head | 4 | 5.6 |
| Thorax | 3 | 4.2 |
| Neck | 1 | 1.4 |
| Total | 72 | 100.0 |

*AIS= Abbreviated injury scale*

**Table IV:** Severity of sports injuries treated in the DGH and DLH during the period of study.

| Variable | Frequency (n=72) | Percentage (%) |
| --- | --- | --- |
| **AIS Score** |  |  |
| 3 Serious | 49 | 68.1 |
| 2 Moderate | 19 | 26.4 |
| 4 Severe | 4 | 5.6 |
| Total | 72 | 100.0 |
| **Play time-loss** |  |  |
| Serious (Over 21 days – permanent) | 50 | 69.4 |
| Moderate (8-21days) | 17 | 23.6 |
| Minor (1-7days) | 5 | 6.9 |
| Total | 72 | 100.0 |

69.4% of injuries resulted to a play time-loss of over 21 days and overall 73.7% of injuries had a score of greater than or equal to three thus were considered to be severe injuries.

## Discussion

This study set out to describe the epidemiological and clinical profile of sports injuries treated at two hospitals in Douala. We assessed 10,211 trauma related files and 114 were sports injuries giving a hospital prevalence of 1.12% amongst all trauma cases. This is similar to the findings of Ibraheem et al who found 0.4% sports injury prevalence amongst all injuries in a teaching hospital in Nigeria [11]. Also Blankson et al found a prevalence of 0.6% sports injuries attending a teaching hospital in Ghana [12]. However Spencer et al found a hospital prevalence of 7.1% in their study. This high prevalence may be linked to the higher hospital access rate and also a higher rate of participation in sports compared to the African context.

The sex distribution showed a male predominance with 86.1% of sports injuries treated in hospitals to have occurred in men. This is in accordance with the findings of Akkaya et al who had 80.5% males [8] and Spencer et al who had 82% males [7]. Sallis et al rather found in their study that female athletes had more injuries 52.5% participant-years than their male counterparts 47.7% participant-years. However, even though this difference was not statistically significant; their study was purely a comparative study in a population strictly made of male and female athletes [13]. This male predominance in our results is due to the high participation of males to females in sports and physical activity in the Cameroonian context [14].

The modal age group was 11-21 with 31.9%, the mean age was 24.97±13.8 years. This result is consistent with the findings of Akkaya et al who found their mean age to be 25.7 ±5.1 years and Spencer et al who had a mean age of 22 years. This similarity could be explained by the high bone growth rate during this age, making bone strength weaker with respect to muscles, tendons and ligaments. This high bone growth rate phenomenon explains why growth plate fractures (weaker zones) and avulsion fractures are more common in children, adolescents and young adults. Also this results may be explained by the very high participation of this age group in recreational sports which have lesser rules and exigencies as concerns protective sports equipment, warm ups, play surfaces etc. The ages ranged from 3 to 69 years old similar to the results of Spencer et al who had an age range from 8 to 60 years, and consistent with the study published by Akkaya et al who found 5 to 68 years. This similarity in age ranges can be explained by the fact that sports injuries affects all age groups.

Students were found with a greater proportion of injuries 48.6%, the employed had 36.1%, 6.9% were unemployed and 8.3% were athletes. This is similar to the findings of Spencer et al who reported in their study that 40.7% were school children or students, 37.2% were employed and 22.1% were unemployed. This similarity could be explained the high participation of students in sporting activities coupled with the pathogenesis related to high bone growth rates in adolescents.

59.7% of injuries occurred during recreational sports then 22.2% occurred during physical education while 11.1% occurred training and 6.9% during competitions. These findings may be explained by the fact that recreational sports do not have exigencies on play surfaces or protective equipment and sometimes lack strict rules. Also most of amateurs and professional athletes attend hospitals less because they usually have an attending physician or onsite medical officer who handles injuries and refer only the serious cases to hospitals.

88.9% of injured persons were non-athletic regular persons, 9.7% amateurs and 1.4% professionals. These findings are similar to Spencer et al who found 62.2% individuals reporting at an accident and emergency department for sports injuries to be non-athletic regular persons, 36.6% were amateurs while only 2% of injuries that attended the emergency department to have occurred to professionals. This similar results can be explained by the fact that these studies are hospital based studies. Also, amateur and professional sporting activities usually have an attending physician who refers only the severe cases for emergency care.

Sport or exercise practiced during which the injury occurred was football in 66.7% cases, running in 13.9% and jumping 12.5% while basketball injuries accounted for 2.8%. This is similar to the findings of Akkaya et al who found football to be the most cause of injuries in all age groups and Spencer et al who found football in 65.11% of cases. This could be explained by the combination of high speed and contact involved in football [15].

The lower extremity was found to be the most injured region most with 47.2% injuries, the upper extremity 29.2%, the face 6.9%, abdomen and pelvis 5.6%, 4.2% of injuries were thoracic. This findings are similar to the study published by Spencer et al showing the majority of injuries were to the extremities (32.6% arm and 55.2% leg), 1.7% chest, 9.3% to head or face, and 1.2% spine. Also, Akkaya et al found similar results in their study with majority of injuries to have occurred in the lower extremity 62.7%, the upper extremity 23.3%, torso 4.7% and the 6.9% were on the head and neck. Systema et al explained that extremity injuries are more common in team sports [16]. Football had the majority injuries, this therefore explains why the majority of injuries in this study occurred to the extremities.

Hard tissue injuries were a majority with 59.7% fractures while 27.7% were soft tissue injuries (8.3% sprains, strains 6.9%, contusion 6.9%, laceration 5.6%) and head injuries 5.6%. This study is not in accord with the findings of Akkaya et al who rather found soft tissue injuries to be a majority of the cases with contusions, lacerations, abrasions and hematoma accounting for 40.6%, strains and sprains 37.5%, while fractures were 15.2% and head injuries 6.5%. Spencer et al reported in their study that majority of the injuries were soft tissue injuries 70.9%, fractures were 17.44% while head injuries 3.5%. This difference in result can be explained by the fact these two hospitals are reference hospitals and also less people access hospitals for soft tissue injuries in Cameroon preferring home massage remedies.

### Limitations

It was a hospital-based, two center limited study and thus is not truly representative of the entire population of Douala, especially since these were tertiary care centers which is not accessible to a large proportion of the population. The study was hospital based meanwhile most sports injuries are community based. Poor storage of file records greatly reduced our study population.

### Research in Context

There are several published studies describing sports related injuries, though most of these were done in high-resource settings with very well organized competitions at the amateur and professional levels. Also, these studies were mostly carried out in community settings whereas a portion sports injuries are treated in hospitals. Given that sports injuries treated in hospitals are career determining injuries which occur in all regions of the world and touch both the pediatric and adult populations, available research has shown interest mostly on developed countries while few or no studies have being conducted in low resource settings precisely sub-Saharan Africa. To get the evidence needed, the authors searched the PubMed and Google Scholar databases, where search criteria included all studies which described sports injuries treated at hospitals or reporting to accident and emergency departments. The Pan African Medical Journal, the Plos One Journal, the British Journal of Sports Medicine, the American Journal of Sports Medicine the International Journal of Sports Medicine were then searched for full articles. Evidence found described the epidemiology of injuries, types of sports practiced at time of injury of body region involved. However, the authors carried out this research in high resource settings leaving out other aspects like injury mechanism, type of injury and severity of injury thus the need for a more thorough investigation in lower resource settings.

The authors feel that this study added the following contributions to the existing information:

- We brought out the hospital prevalence of sports injuries treated at low resource setting hospitals.
- We described epidemiological findings as concerns the profile of injured patients and the sport done at time of injury in a low –income setting.
- We outlined the type of sports injuries treated at low resource setting hospitals.
- We outlined the percentages of injuries that occurred on different body segments in lower resource settings.
- We outlined the severity of sports injuries treated in lower resource setting hospitals.

Combined with the existing information, this study has the potential to provide sport medicine physicians, clinicians, coaches and athletes in low-income settings with information on the prevalence and severity of sports injuries in low resource settings. This information has been described in the full study (available at the library of the Faculty of Medicine and Pharmaceutical Sciences, University of Douala, the library of the Douala General Hospital and Douala Laquintinie Hospital). This information has the potential to reduce injury severity by increasing injury prevention in the risk groups and emphasizing on the severity of injuries treated at hospitals thus reducing sports injuries in low-resource settings. This information will also increase awareness amongst clinicians, orthopedic surgeons and sports medicine physicians in low resource settings and the world at large to devise strategies in the prevention and treatment of sports injuries.

## Data Availability

All relevant data are within the manuscript and its Supporting Information files. Supporting information files will be uploaded as requested.

